# A novel sample pooling strategy and its application for mass screening of SARS-CoV-2 in an outbreak of COVID-19 in Vietnam

**DOI:** 10.1101/2020.09.11.20192484

**Authors:** Ton That Thanh, Nguyen Thi Thanh Nhan, Huynh Kim Mai, Nguyen Bao Trieu, Le Xuan Huy, Ho Thi Thanh Thuy, Le Thanh Chung, Nguyen Ngoc Anh, Nguyen Thi Thu Hong, Bui Duc Thang, Nguyen Thi Hoai Thu, Le Thi Kim Chi, Nguyen Thi Hanh, Nguyen Huy Hoang, Nguyen Van Vinh Chau, Guy Thwaites, Do Thai Hung, Le Van Tan, Ngo Thi Kim Yen

## Abstract

We present a sample pooling approach and the results of its application for mass screening of SARS-CoV-2 in >96,000 asymptomatic individuals. Our approach did not compromise the sensitivity of PCR, while increasing the throughput and reducing 77% of the costs. 22/32 asymptomatic cases would have been missed without mass screening.

---

Severe acute respiratory syndrome coronavirus 2 (SARS-CoV-2) is the cause of coronavirus disease 2019 (COVID-19) pandemic [1]. As of 11 September 2020, there have been >28 million cases and >913,000 deaths reported globally (https://who.int). After the first infection reported in Vietnam on 23 January 2020 [2], COVID-19 was brought under control by the second week of April [3]. However, after 100 days of no community transmission, on 25 July 2020, SARS-CoV-2 infection was confirmed in a 57 year-old man presenting with pneumonia admitted to C Hospital in Da Nang city, central of Vietnam. This was followed by the detection of SARS-CoV-2 in another 61-year old man who had been mechanically ventilated for four days in another hospital of the city, Da Nang Hospital [4].

Rigorous contact tracing and testing resulted in an escalation of COVID-19 cases in the following week, with the majority of cases being linked with Da Nang hospital [4]. Consequently, the city was locked down on July 28, 2020. In parallel, a mass community testing approach coupled with a novel sample pooling strategy was implemented on the second week of August 2020, contributing to the success of COVID-19 control in Da Nang city. Indeed, as of 11 September 2020, the city experienced 12 consecutive days with no community transmission [4]. Here, we describe our sample pooling strategy and the results of its application for mass community testing for the period between 8 and 21 August 2020.

## METHODS

### COVID-19 containment approaches applied in Da Nang

In addition to the containment approaches widely applied in Vietnam [5], a mass community testing was implemented in Da Nang from the second week of August 2020 onward. Accordingly, this approach has been applicable to asymptomatic individuals with at least one of the criteria: 1) quarantined individuals with the first SARS-CoV-2 RT-PCR negative, 2) visiting Da Nang Hospital between 15 and 26 July 2020, the period with ongoing transmission within the hospital as defined by the local authorities, 3) living in an area where there was a confirmed case of community transmission, 4) a family member of person who was in direct contact of a confirmed case.

### Collection of pooled nasal pharyngeal throat swabs and epidemiological data

We pragmatically aimed to group 2-7 people into one group for testing. We collected a nasal pharyngeal throat swab (NTS) from each individual of the group, and combined in a single 15-ml collection tube containing 3 ml of viral transport medium (Figure 1A). If SARS-CoV-2 testing of the pool returned positive, we then collected single NTS from each individuals of the corresponding group for confirmatory RT-PCR testing (Figure 1A). All collected samples were sent to the laboratory of Da Nang Centre for Disease Control and Prevention which was responsible for 90% of mass screening in Da Nang city. We also collected demographics, contact history and respiratory signs/symptoms from the confirmed cases using a standard case record form.

**Figure 1A:**
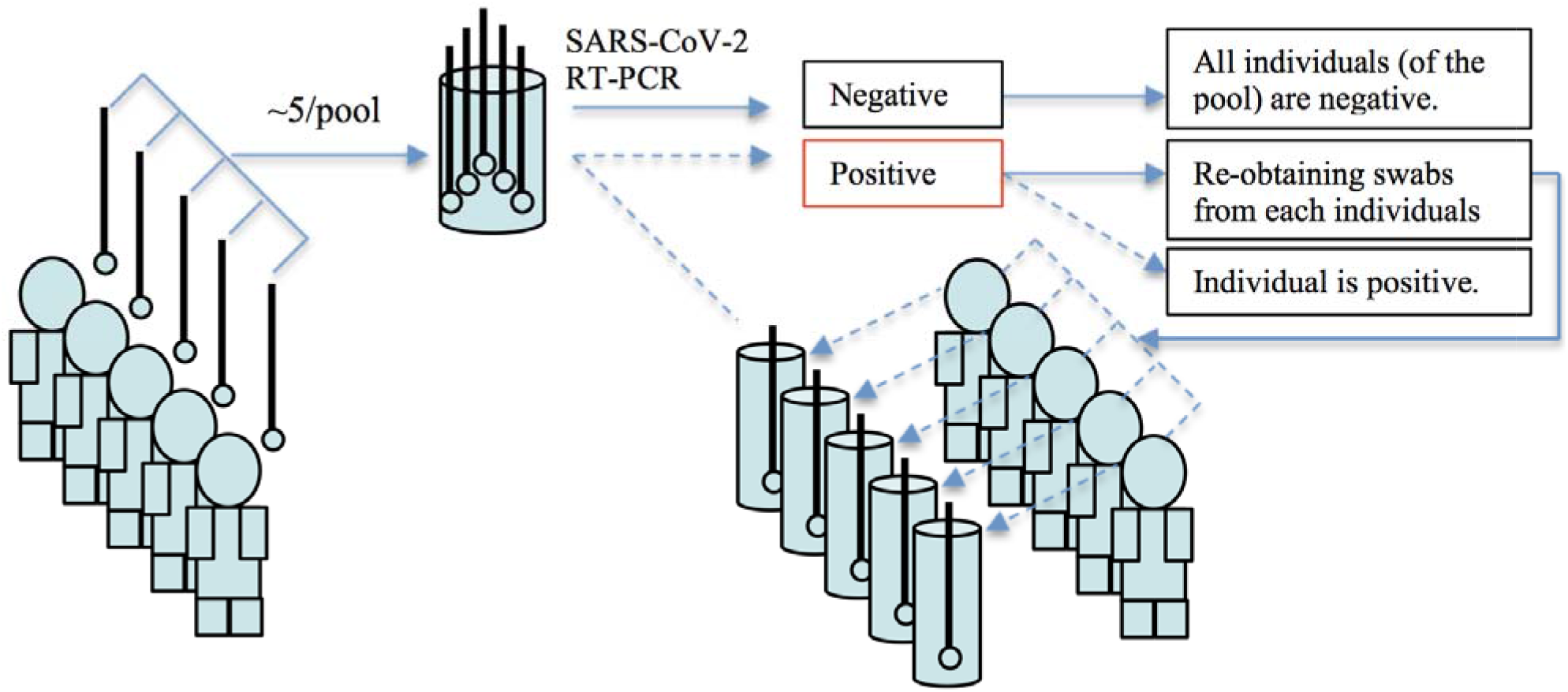
Flowchart illustrating the strategy of obtaining the pooled nasal pharyngeal throat swabs for primary screening of SARS-CoV-2, and follow-up on individuals of the pools that had a positive SARS-CoV-2 RT-PCR result.

### Viral RNA extraction and SARS-CoV-2 RT-PCR

We extracted viral RNA from 200ul of the pooled NTS samples using Thermo Scientific KingFisher Flex, an Automatic Nucleic Acid Extraction Workstation (Thermo Scientific), following manufacturer’s instructions. We used LightPower iVASARS-CoV-2 1st RT-rPCR Kit (Viet A, Vietnam) as a screening assay, and the E gene real time RT-PCR assay (TIB MOLBIOL, Germany)[6] as a confirmatory assay.

For subsequent testing of NTS swabs obtained from individuals of the corresponding positive pools (Figure 1A), we manually extract viral RNA from 140ul of the NTS samples using the QIAamp viral RNA kit (QIAgen GmbH, Hilden, Germany), following the manufacturer’s instruction, and used the TIB MOLBIOL E gene real time RT-PCR to detect SARS-CoV-2.

### Statistical analysis

We used Wilcoxon signed-rank text available in GraphPad Prism version 5.04 (GraphPad Software, San Diego, California) to compare the Ct values obtained from the pooled samples and individual samples. The Ct values used for the analysis were those generated by the confirmatory assay.

### Ethics

The present work formed part of the response to COVID-19 outbreaks that was approved by Da Nang Department of Health. Accordingly, obtaining inform consent from individuals was deemed unnecessary.

## RESULTS

### Collection of pooled nasal pharyngeal throat swabs

From August 8 to August 21, 2020, a total of 22,290 pooled NTS samples were collected from 96,123 individuals meeting the inclusion, and successfully tested for SARS-CoV-2. The number of NTSs per pool varied between 2 and 7 with the majority having 5 samples per pool (Supplementary Table 1). The included individuals accounted for 8.2% of the population of 1.1 million in Da Nang city, and came from 7/8 eight districts of the city. The remaining district had no reported COVID-19 case as of September 5, 2020.

### SARS-CoV-2 detection in pooled/individuals nasal pharyngeal throat swabs

RT-PCR testing revealed evidence of SARS-CoV-2 RNA in 24/2290 (0.11%) of the pools (Supplementary Table 1), with Ct values of internal controls within the normal range (data not shown). Consequently, a total of 104 individuals were resampled and tested for SARS-CoV-2 by RT-PCR. Subsequently, 32 individuals belonging to one of the 24 originally positive pools had a confirmed diagnosis of SARS-CoV-2 (Figure 1B), accounting for 21% reported cases in Da Nang City during the same period (8-22 August 2020) (Supplementary Figure 1). There was no difference in Ct values of pooled NTS samples and that of individual NTS samples (Figure 1C).

**Figure 1B:**
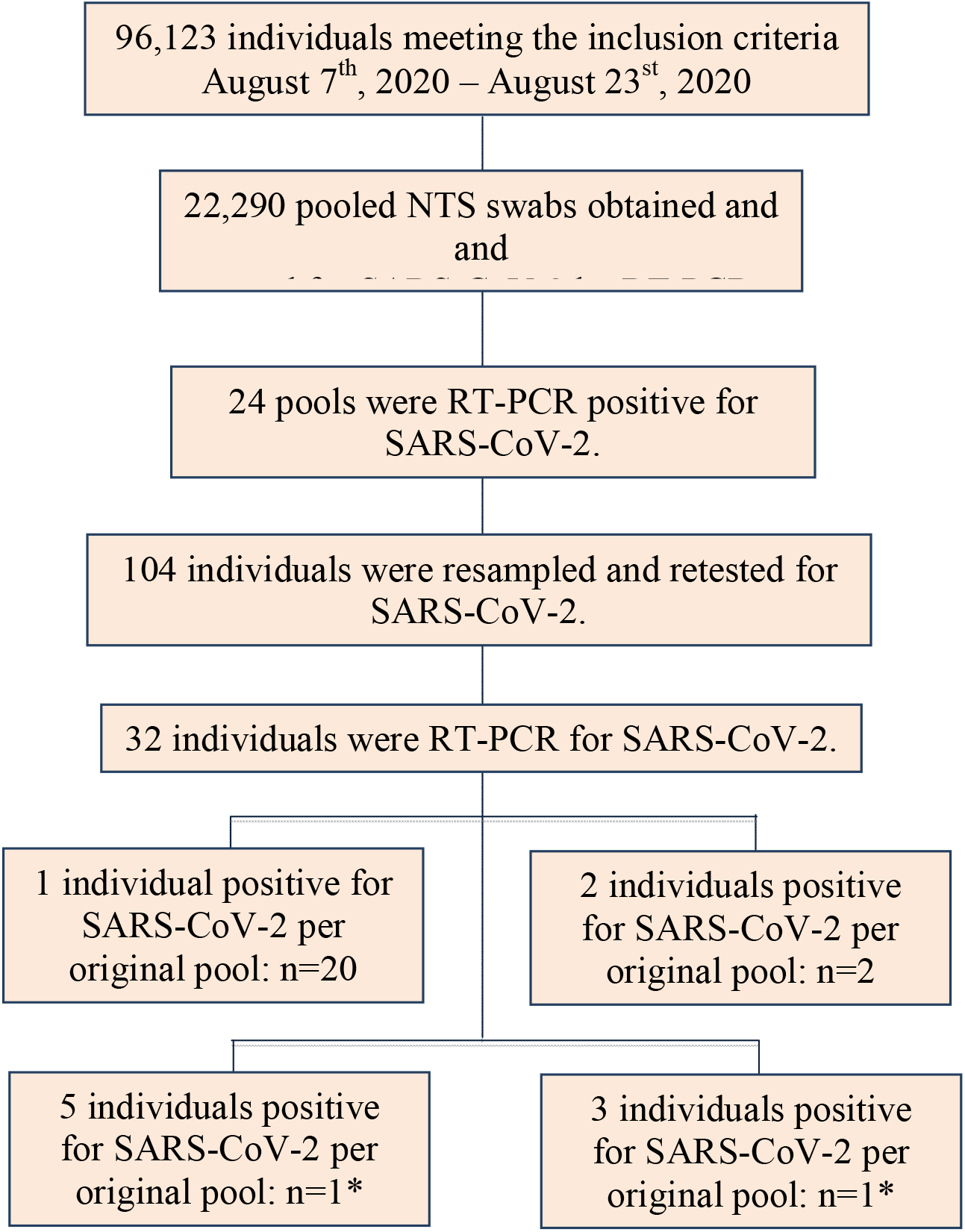
Flowchart illustrating the mass community testing for SARS-CoV-2 leading to the detection of 32 cases, including eight belonging to two family clusters **Note to Figure 1B**: *From the same family of five and three people, respectively who were all positive for SARS-CoV-2, but were all asymptomatic.

**Figure 1C:**
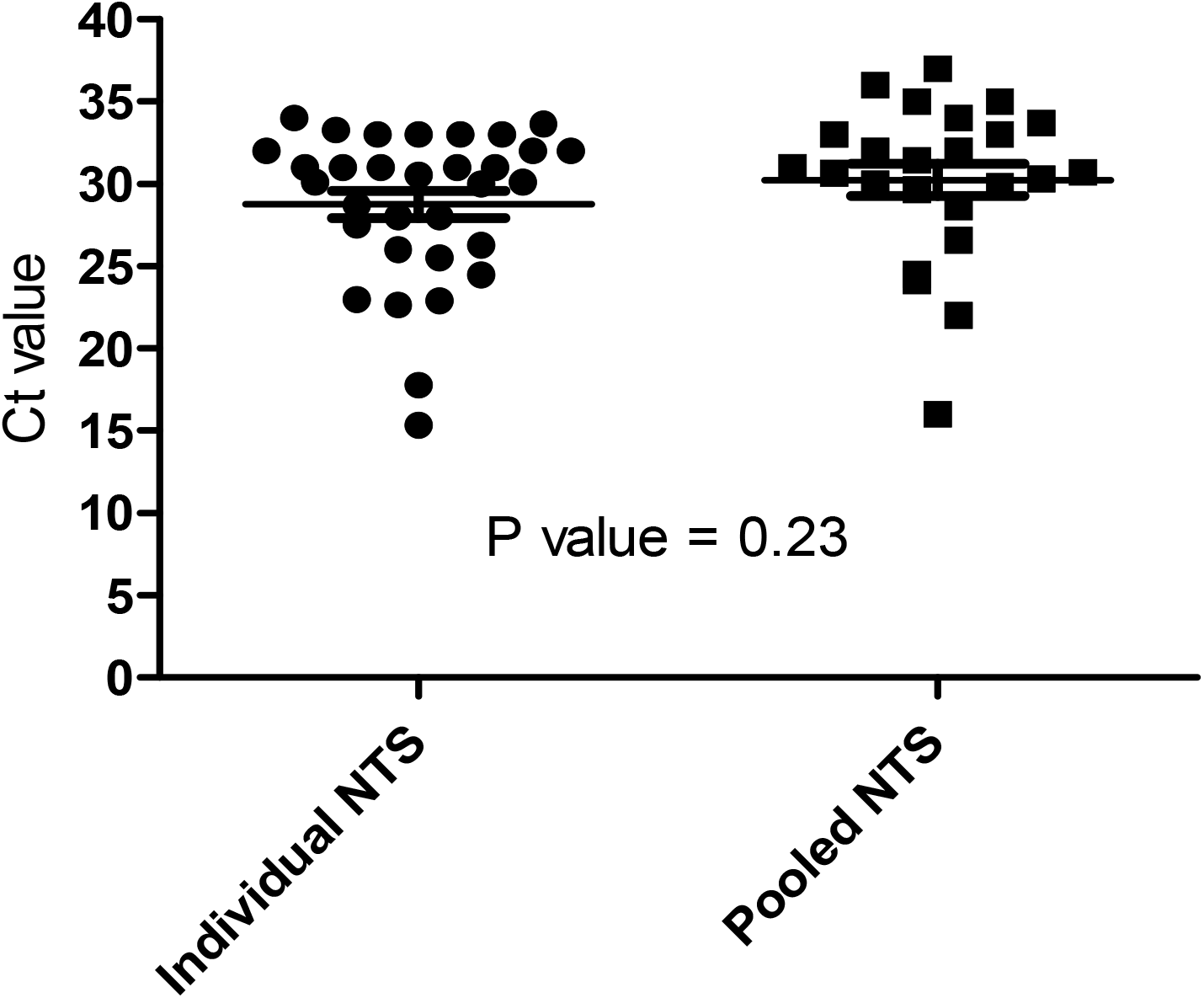
Plots showing real time RT-PCR Ct values of pooled NTS samples and individual NTS swabs **Note to Figure 1C**: a total of 24 pooled NTS samples and 32 individual NTS samples were included for analysis. Ct values (median, range): for pooled NTS samples: 30.86 (16 – 37), for individual NTS samples: 30.33 (15.36 – 34). P comparison = 0.23

The time from pooled sample collection to reporting the confirmed diagnostic result was either the same day (n=9), 24 hours (n=10) or 48 hours (n=12). In the remaining case, the result was released within 3 working days.

### Demographics, clinical characteristic and contact history of the confirmed cases

The 32 SARS-CoV-2 positive individuals included 21 females and 11 males aging from 14 to 73 years (median 45 years). None had any signs/symptoms of respiratory infection at the time of collection. Of these, 22 had no history of contact with a confirmed COVID-19 case (Supplementary Table 2 and Figure 1B). Two family clusters with all eight members were positive for SARS-CoV-2 and were all asymptomatic. None recalled of having any signs/symptoms of respiratory infections.

### The effectiveness of group testing strategy

By pooling, we successfully saved 73,833 RT-PCR tests and associated consumables (corresponding to a reduction of 77% of the costs). In our setting, the associated cost to conduct RT-PCR testing for SARS-CoV-2 in one suspected case was ∼21.5 US$. Thus, excluding manpower, our mass screening strategies saved ∼1.5 million US$. Because our available resources could only accommodate some ∼1500 SARS-CoV-2 RT-PCRs per days for mass screening, it would have taken 64 days to complete the screening for those 96,123 individuals compared to 14 days required by our pooling strategy.

## DISCUSSION

We present a novel sample pooling strategy and the results of its application for mass community screening of SARS-CoV-2 in asymptomatic people in Da Nang city, an epicenter of the ongoing COVID-19 outbreak in Vietnam since the end of July 2020. We tested 96,123 asymptomatic individuals and found 32 confirmed cases (0.03%); accounting for 21% of 156 reported cases in Da Nang city during the same period. Of these, 22 had no history of contact with a confirmed case. Therefore, they would have been missed, should mass community testing had not been implemented, and may have contributed to the expansion of the outbreak.

In previous reports, individual samples were separately collected, and an equal volume from several samples was then pooled into one tube prior to nucleic acid isolation for subsequent RT-PCR analysis [7, 8]. This dilutes individual samples of the pools, and therefore could in principle compromise the sensitivity of the RT-PCR [7, 9], especially in case of samples with low viral loads [10]. In contrast, our strategy combined individual swabs in one tube at the time of collection, therefore did not result in dilution of individual samples. Indeed, we observed no difference in the obtained Ct values between the pooled and individual NTS samples.

Potentially, the reduction in time required to complete the screening of 96,123 individuals from 64 days to 14 days has had significant implication for the success of COVID-19 control in Da Nang to date. In addition, combining individual swabs at collection reduced 77% of the laboratory spaces required to accommodate all NTS samples from those 96,123 individuals, and substantially reduced the reagents and manpower associated costs. These costs and time savings are critical to COVID-19 control, especially in resource limited settings.

In spite of these advantages, confirmatory testing of samples obtained on the other days after the primary screening might result in false negative results, especially during the later phase of the infection [8]. We were not able to assess this in our study, although at least one sample from all 32 positive pools was subsequently positive for SARS-CoV-2.

To summarize, we showed that combining NTS samples at the time of collection allows for high throughput mass screening of asymptomatic individuals, while maintaining the sensitivity and substantially reducing the associated costs. The approach might be applicable in other settings, where there are shortages of diagnostic reagents and the disease prevalence is low, but the demands for testing is high.

## Data Availability

All data are presented in the ms.

## ACKNOWLEDGEMENTS

The study was funded by Da Nang People’s Committee, Da Nang, Vietnam. We would like to thank the World Health Organization and the US Centers for Disease Control and Prevention for their support with the COVID-19 diagnostic reagents. LVT and GT are supported by the Wellcome Trust of Great Britain (204904/Z/16/Z and 106680/B/14/Z, respectively)

---

**Supplementary Table 1:** Details about the number of individuals per pool nasal pharyngeal throat swabs and results of RT-PCR testing for SARS-CoV-2

| Number of individual swabs per pool | Number of pools, N=22,290 | Number of pools with SARS-CoV-2 detected (%) | Number of people positive for SARS-CoV-2 after resampling and testing of individual samples |
| --- | --- | --- | --- |
| 2, n(%) | 1408 (6) | 1 (0.07) | 1 |
| 3, n(%) | 3337 (15) | 4 (0.12) | 6 |
| 4, n(%) | 4528 (20) | 5 (0.11) | 5 |
| 5, n(%) | 12922 (58) | 14 (0.11%) | 20 |
| 6, n(%) | 91 (0.4) | 0 | Non applicable |
| 7, n(%) | 4 (0.02) | 0 | Non applicable |

**Supplementary Table 2:** Epidemiological factors of SARS-CoV-2 positive individuals

| Patient Number* | Reason for being tested | Recalled date of contact | Tested positive on | Risk group<sup>#</sup> |
| --- | --- | --- | --- | --- |
| 1 | A neighbor of a confirmed case. No known contact with a confirmed case |  | August 9 | 1 |
| 2 | Living in the same apartment buildings of a confirmed cases | Non applicable | August 10 | 1 |
| 3 | A neighbor of a confirmed case. No known contact with a confirmed case | Non applicable | August 10 | 1 |
| 4 | Visited Da Nang hospital at any time point July 15, 2020 – July 26, 2020. No known contact with a confirmed case | Non applicable | August 11 | 1 |
| 5 | Visited Da Nang hospital at any time point July 15, 2020 – July 26, 2020. No known contact with a confirmed case | Non applicable | August 11 | 1 |
| 6 | No known contact. Living in an area with recorded community transmission | Non applicable | August 12 | 1 |
| 7 | Visited Da Nang hospital at any time point July 15, 2020 – July 26, 2020. No known contact with a confirmed case | Non applicable | August 12 | 1 |
| 8 | Visited Da Nang hospital at any time point July 15, 2020 – July 26, 2020. No known contact with a confirmed case | Non applicable | August 15 | 1 |
| 9 | Visited Da Nang hospital at any time point July 15, 2020 – July 26, 2020. No known contact with a confirmed case | Non applicable | August 15 | 1 |
| 10 | Visited Da Nang hospital at any time point July 15, 2020 – July 26, 2020. No known contact with a confirmed case | Non applicable | August 15 | 1 |
| 11 | A neighbor of a confirmed case with out a known contact | Non applicable | August 15 | 1 |
| 12 | Visited Da Nang hospital at any time point July 15, 2020 – July 26, 2020. No known contact with a confirmed case | Non applicable | August 17 | 1 |
| 13 | Visited Da Nang hospital at any time point July 15, 2020 – July 26, 2020. No known contact with a confirmed case | Non applicable | August 17 | 1 |
| 14 | A businessperson at a market where mass community testing was conducted. No known contact with a confirmed case | Non applicable | August 18 | 1 |
| 15 | No known contact. Living in an area with recorded community transmission | Non applicable | August 18 | 1 |
| 16 | Visited Da Nang hospital at any time point July 15, 2020 – July 26, 2020. No known contact with a confirmed case | Non applicable | August 20 | 1 |
| 17 | Visited Da Nang hospital at any time point July 15, 2020 – July 26, 2020. No known contact with a confirmed case | Non applicable | August 20 | 1 |
| 18 | Visited Da Nang hospital at any time point July 15, 2020 – July 26, 2020. No known contact with a confirmed case | Non applicable | August 20 | 1 |
| 19 | A businessperson at a market where mass community testing was conducted. No known contact with a confirmed case | Non applicable | August 22 | 1 |
| 20 | A businessperson at a market where mass community testing was conducted. No known contact with a confirmed case | Non applicable | August 21 | 1 |
| 21 | A businessperson at a market where mass community testing was conducted. No known contact with a confirmed case | Non applicable | August 21 | 1 |
| 22 | A guard at a market where mass community testing was conducted. No known contact with a confirmed case | Non applicable | August 22 | 1 |
| 23 | A contact of a confirmed case. First SARS-CoV-2 RT-PCR was negative. | July 20, 2020 | August 12 | 2 |
| 24 | A family member (patient 26) was a contact of a confirmed case. | July 20, 2020 onward | August 12 | 2 |
| 25 | A family member (patient 26) was a contact of a confirmed case. | July 20, 2020 onward | August 12 | 2 |
| 26 | A family member (patient 26) was a contact of a confirmed case. | July 20, 2020 onward | August 12 | 2 |
| 27 | A family member (patient 26) was a contact of a confirmed case. | July 20, 2020 onward | August 12 | 2 |
| 28 | A contact of a confirmed case. First SARS-CoV-2 RT-PCR was negative. | July 20, 2020 onward | August 15 | 2 |
| 29 | A contact of a confirmed case. First SARS-CoV-2 RT-PCR was negative. | July 24, 2020 onward | August 17 | 2 |
| 30 | A contacts of two confirmed cases. First SARS-CoV-2 RT-PCR was negative. | July 25, 2020 onward | August 19 | 2 |
| 31 | A contact of a confirmed case. First SARS-CoV-2 RT-PCR was negative. | August 12, 2020 | August 20 | 2 |
| 32 | A contact of a confirmed case. First SARS-CoV-2 RT-PCR was negative. | July 24, 2020 | August 20 | 2 |

**Note to Supplementary Table 2**: *none presenting with any respiratory sign/symptom at the time of swab collection of mass community testing.^#^1: would have been missed without mass community testing, 2: could be picked up through contact tracing and PCR testing of contacts of a confirmed case.

**Supplementary Figure 1:**
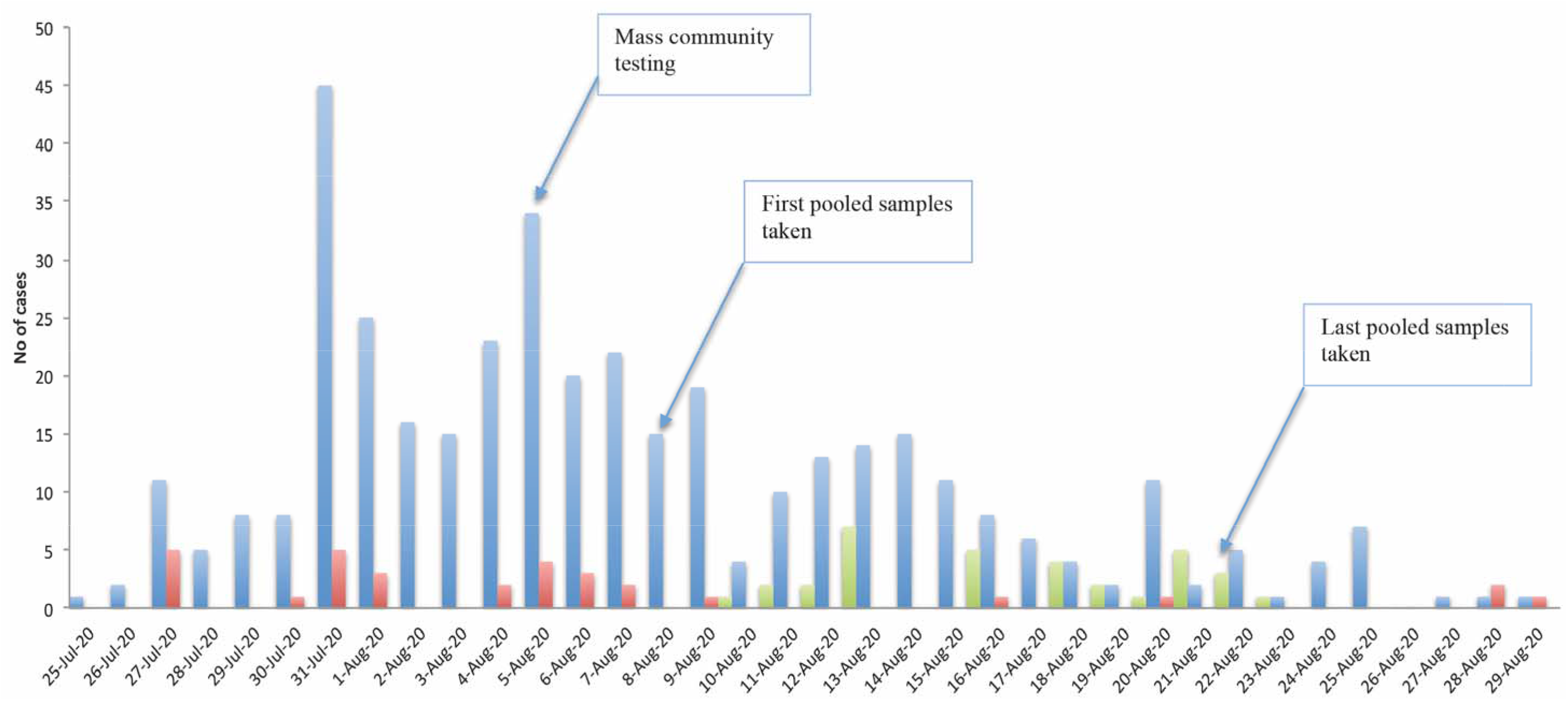
Temporal distribution of the number of reported cases (blue bars) and deaths (red bars) in Da Nang between July 25 and August 29, 2020, and the number of cases detected through the mass community testing approach (green bars)

